# Understanding Malaria Treatment Adherence in Rwanda: Implications for Artemisinin Resistance

**DOI:** 10.1101/2025.07.22.25331991

**Authors:** Pierre Gashema, Aileen Jordan, Eric Saramba, Neeva Wernsman Young, Patrick Gad Iradukunda, Corine Karema, Jean Baptiste Mazarati, Jonathan J Juliano, Jeffrey A. Bailey, Kristin Banek

**Affiliations:** College of Medicine and Veterinary Medicine, University of Edinburgh, Edinburgh, UK; Center for Genomic Biology, INES-Ruhengeri, Ruhengeri, Rwanda; College of Medicine and Health Sciences, University of Rwanda, Kigali, Rwanda; Center for Computational Molecular Biology, Brown University, Providence RI, USA; Repolicy Research Centre, Kigali, Rwanda; Drug Department, Rwanda Food and Drugs Authority, Kigali, Rwanda; Quality Equity Health Care, Kigali, Rwanda; Division of Infectious Diseases, School of Medicine, University of North Carolina, Chapel Hill, NC, USA; Department of Pathology, Brown University, Providence, RI, USA; Institute for Global Health and Infectious Diseases, University of North Carolina, Chapel Hill, NC, USA; Department of Epidemiology, Tulane University Celia Scott Weatherhead School of Public Health and Tropical Medicine, New Orleans, LA

**Keywords:** Malaria, medication adherence, knowledge, attitudes, and practices (KAP), Artemisinin resistance

## Abstract

Prompt diagnosis and effective treatment are key malaria interventions that rely on community knowledge and adherence to treatment. With the emergence of artemisinin resistance in Rwanda, ensuring optimal malaria treatment practices within communities is essential. This study examined malaria knowledge, attitudes, and practices (KAP) among febrile patients at government clinics to identify factors affecting influencing malaria treatment practices. A cross-sectional study was conducted in six health facilities in moderate to high malaria transmission areas of Rwanda. Patients or caregivers of children with fever were enrolled and interviewed using semi-structured questionnaires. From December 2023 to February 2024, 406 participants were enrolled, 56% (228/406) were female, 50% (204/406) were primary schooled, and mostly in rural areas 80% (324/406). A total of 71% (289/406) of participants owned insecticide-treated nets (ITNs), and 51% (205/406) received indoor residual spraying (IRS). Malaria knowledge was high among respondents, with 81% (329/406) correctly identifying symptoms, 72% (291/406) understanding transmission modes, and 74.6% (303/406) aware of effective control measures. However, of the 44.3% (180/406) who received malaria treatment in the last 6 months, only 46% (83/180) completed the appropriate 3-day medication course; 37% (66/180) stopped within 2 days, and 11% (19/180) over 3 days. Furthermore, 27% (109/406) of participants took antimalarials for fever; the majority (54%; 49/109) received medication from drug outlets/pharmacies. Although knowledge and attitudes toward malaria treatment were high, adherence was poor, thereby exacerbating the risk of developing resistance. Effective interventions are urgently needed to improve antimalarial adherence, particularly in sub-Saharan African countries with documented antimalarial resistance.

## Introduction

Despite significant gains, malaria remains a major public health concern, with the World Health Organization (WHO) reporting 249 million cases in 2022, an increase of five million cases observed since 2021^1^ The vast majority of malaria-related mortality (95.4%) remains in the WHO Africa region.^1^ Over the past two decades, Rwanda has seen significant variation in malaria incidence. During the period 2005-2010, malaria incidence in Rwanda decreased from 162 per 1000 population to 62 per 1000, with the dramatic decrease linked to multiple malaria control interventions (i.e., improved treatment and vector control)^2^ as well as the adoption of artemisinin-based combination therapies (ACTs) as the first-line malaria treatment in 2006^3^. These interventions resulted in a decrease of over 50% in associated admissions and deaths at health facilities during this time period.^3^ However, between 2011 and 2017, the Rwanda National Malaria Control Program (NMCP) reported an eightfold increase in malaria cases countrywide from 45 to 486 per 1000 population.^4^ This resurgence was likely driven by multiple factors, both related and unrelated to specific malaria control interventions.^5^

To combat this resurgence, the Rwanda Malaria Strategic Plan 2020-2024 aimed to reduce mortality and morbidity related to malaria by employing key malaria control interventions.^6^ Besides vector control interventions, malaria control depends on effective case management, which requires timely, accurate diagnosis and, importantly, appropriate treatment with effective antimalarials.^2^ In part, the NMCP instituted the expansion of malaria treatment from central to peripheral levels, including community-based case management with approximately 60,000 Community Health Workers (CHWs) (two per village referred to as binômes).^5^ CHWs are trained through complementary programs of Integrated Community Case Management of Fever (iCCM) and Home-Based Management of Fever (HBM) to use malaria rapid diagnostic tests (RDTs), dispense ACTs, and refer severe cases.^7^ The integration of CHWs helped the Rwanda Ministry of Health to respond promptly to malaria cases, and according to WHO, 55% of malaria cases were diagnosed and treated by CHWs in Rwanda by the end of 2022 compared to 15% in 2016.^8^ However, artemisinin partial resistance (ArtR) threatens this new progress. Rwanda was the first African nation to document ArtR in cases from 2014,^9^ and now ArtR conferring mutations are prevalent nationwide, complicating the future of malaria treatment in Rwanda. ^10–12^

The overarching goal of the national malaria strategic plan is to interrupt malaria transmission.^13^ Community education and knowledge about malaria is important for effective resource utilization and will likely become more important as malaria transmission decreases, resulting in less daily contact with the disease.^14^ Rwanda has assessed community knowledge and practice on malaria interventions, however the data is limited. In Ruhuha, one of the endemic areas in Eastern Rwanda, a survey conducted in 2015 showed that 80.1%, 91.4%, and 87.3% of the community had good knowledge, attitudes, and practices towards malaria interventions.^15^ In 2015, Asingizwe et al. evaluated Rwandan community readiness on pre-elimination in the same area and revealed that 90.7% of individuals were knowledgeable about malaria elimination strategies.^16^ In neighboring Uganda, local knowledge of malaria among patients was shown to drive appropriate case management and use of antimalarials.^17^ To update and assess more broadly across Rwanda, community-level knowledge and potential factors influencing malaria treatment adherence behaviors, we conducted a malaria Knowledge, Attitude, and Practices (KAP) study among patients seeking treatment for fever at six clinics across Rwanda to identify factors associated with malaria treatment practices.

## Methodology

### Study Context

This study was conducted in six governmental health facilities (Ndama and Mahama in the Eastern Province, Mushubati in the Western Province, Tanda in the Northern Province, Gishubi in the Southern Province, and Masaka in Kigali City) across rural and urban areas in Rwanda. We deliberately selected these health facilities based on case load, endemicity, and geographical context (urban/rural), ensuring representation from all provinces (**Figure 1**).^18^ A convenience sampling approach was used, where febrile patients were recruited after their consultations. Eligible participants provided informed consent and completed a structured questionnaire, with each interview lasting approximately 30 minutes. After each interview, the next patient exiting the consultation room was screened, ensuring they had fever but no severe disease or other conditions preventing participation. This sequential process ensured systematic recruitment while reducing selection bias.

**Figure 1.**
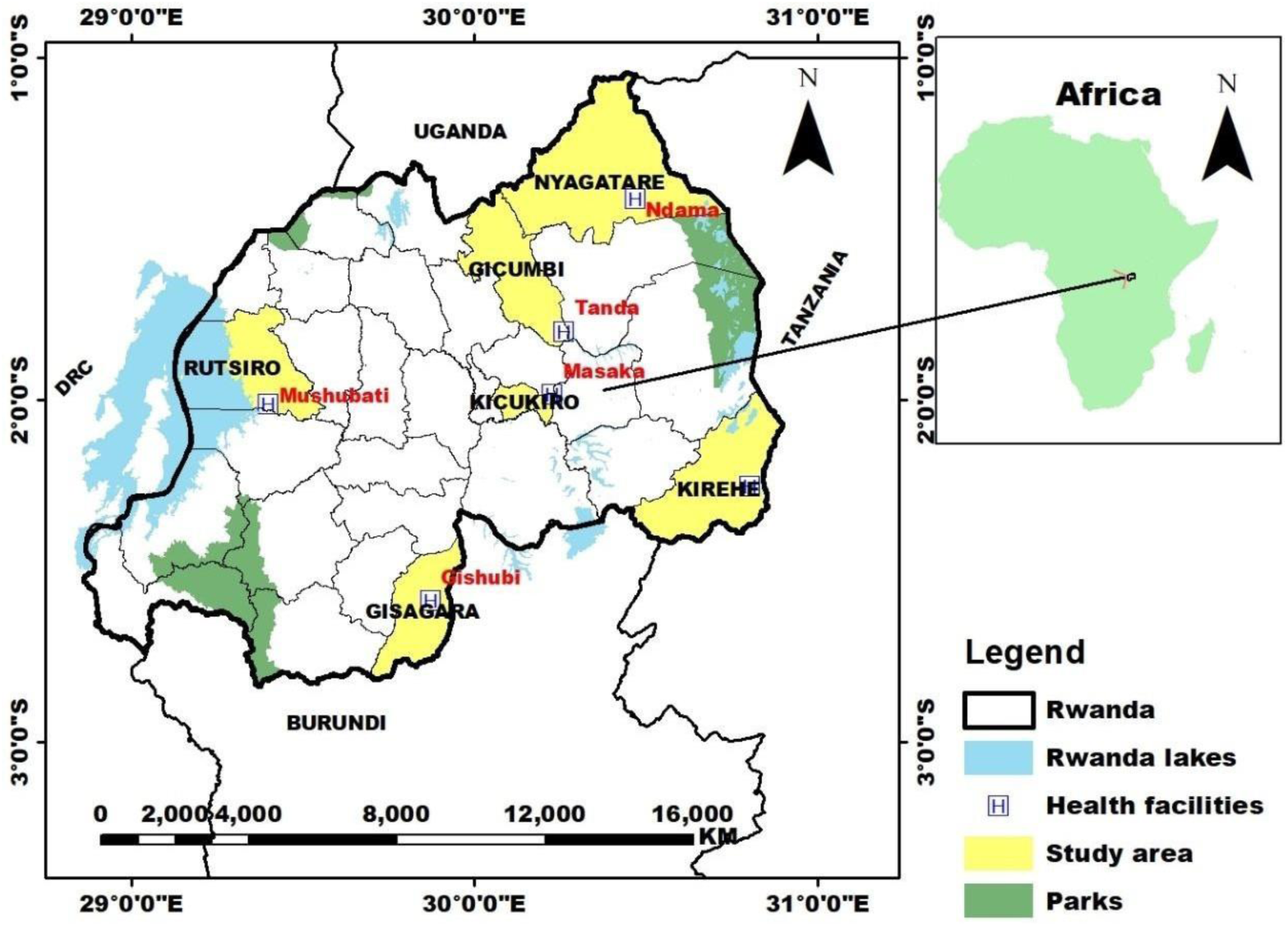
Map of the study area. Health Facilities (red) and their corresponding districts (yellow) where the study was conducted. Kirehe, Gisagara, and Nyagatare are considered high malaria transmission areas, whereas Kicukiro, Mushubati, and Gicumbi are classified as moderate transmission areas. Except for Kicukiro District, the others are rural areas.

### Study Design and Sample Size

A health facility-based cross-sectional study was conducted to assess the knowledge, attitudes, and practices regarding malaria treatment among patients seeking treatment for fever. The aim was to identify factors that influence malaria treatment practices. The intended sample size of 400 fever patients and was calculated using Yamane’s formula based on an estimated population of 13 million for Rwanda, a margin of error (*e*) of 0.05.^19^

### Data Collection

A standard KAP questionnaire was developed based on previous Rwanda Malaria Indicator Surveys and the National Malaria Survey in Sierra Leone.^18,20^ The questionnaire included demographic characteristics of the participants, knowledge of participants on malaria disease (symptoms and transmission), preventive measures for malaria, malaria treatment, and attitude toward existing control measures and treatment practices as shown in supplementary file 1 (**S1**). All questionnaires and consent forms were first developed in English and later translated into Kinyarwanda and back to English to ensure that meaning was retained. Each participant was assigned a unique identification number based on the name of the health facility.

### KAP Scores and Outcome Definitions

Quality scores were derived to assess participants’ knowledge, attitudes, and practices. The score for each participant was obtained by summing the scores of all items in the three sections and then calculating the total as a percentage of the maximum possible score. The scaled quality score is calculated using the formula: Scaled Quality Score (Obtained Score minus (-) Minimum Possible score divided by Maximum possible score minus (-) Minimum Possible Score) × 100%.^21^ Treatment adherence was defined as taking antimalarial treatment for three days.

### Data Management and Analysis

Responses were collected using paper forms, and all data were entered and validated in a Microsoft Excel spreadsheet. Analyses were conducted in SPSS (version 2023) (IBM, New York, USA) and STATA 16 (StataCorp, LLC, College Station, TX). Basic descriptive statistics were used to calculate the frequencies and proportions of demographic characteristics of participants and the KAP indicators. Logistic regression was used to estimate the crude (bivariate) and adjusted (multivariate) odds ratios and their 95% confidence intervals to assess the association between participant characteristics and KAP indicators and the two adherence outcomes (diagnosed malaria treatment and fever treatment. Covariates were assessed for correlation using Pearson’s correlation coefficient; none were found to be strongly correlated (r ≥ 0.8). All covariates were included in multivariable analyses regardless of p-values. Covariates with small cell sizes or low frequencies were excluded from multivariable analysis to avoid bias and unreliable estimates. Associations were considered significant if the p-value was < 0.05.

### Ethical Considerations

The protocol was reviewed and approved by the University of Edinburgh Medical School and the Rwanda National Ethics Committee (RNEC) with approval number (Ref No. RNEC 227/2023). Written informed consent was obtained from participants before participating in the study.

## Results

### Participant Characteristics

A total of 406 patients/caregivers were enrolled in the study from December 2023 to February 2024. Of those enrolled, 17% (69/406) of them were from Gishubi Health Center, 16% (66/406) from Mahama, 17% (70/406) from Masaka, 16% (65/406) from Mushubati, 18% (68/406) Ndama and17% (68/406) Tanda. Demographic characteristics are summarized in **Table 1**. Over half 56% (228/406) of participants were female, and 80% (324/406) lived in rural areas. Half of all respondents attended primary school, and 28.8% (88/406) had the secondary level of education and above. The mean age of the study participants was 29. The majority of participants 59.6%(242/406) were farmers. The primary religion of participants was Catholic 40.1% (163/406).

**Table 1.**
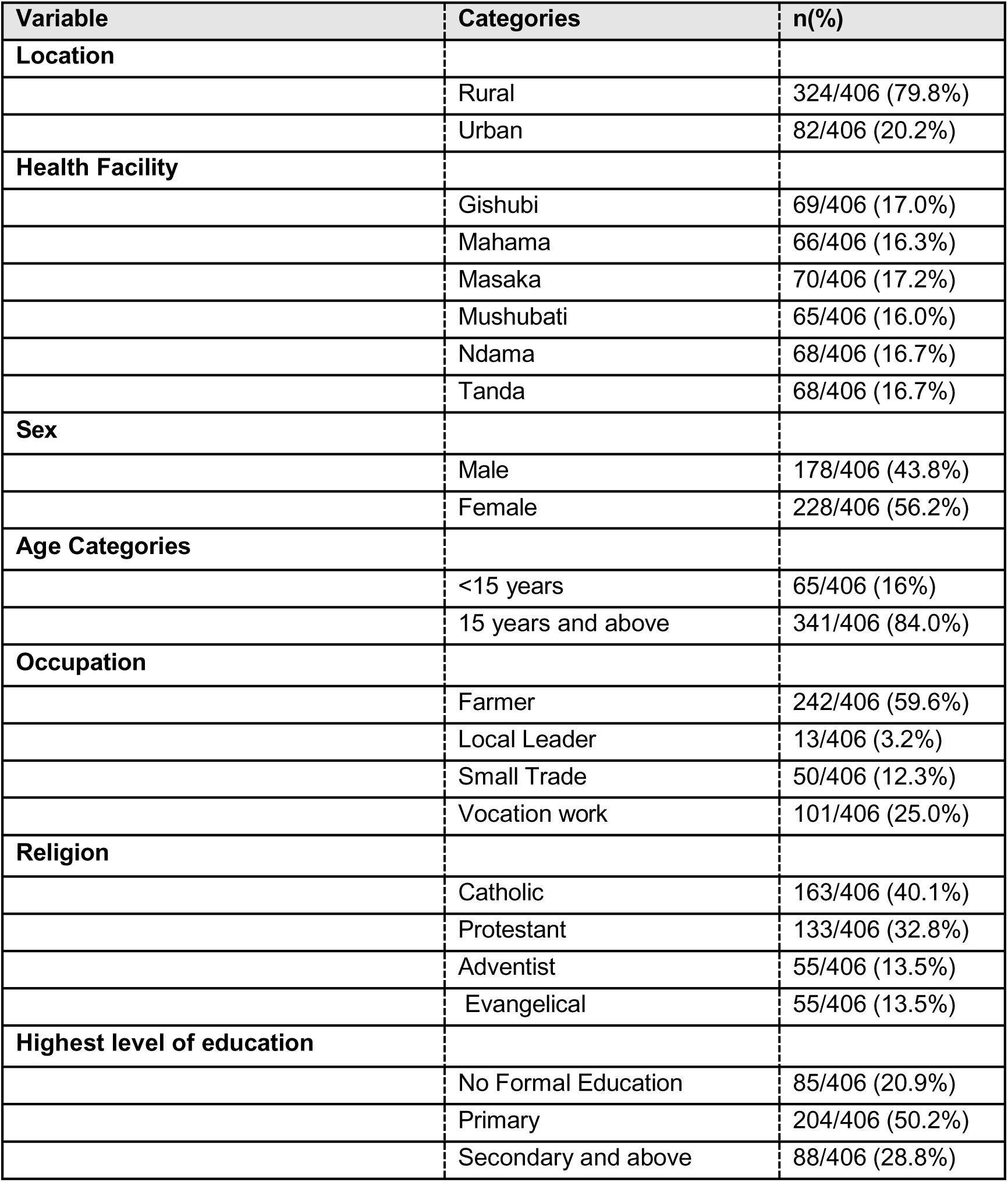
Participant Characteristics (n=406)

| Variable | Categories | n(%) |
| --- | --- | --- |
| **Location** |  |  |
|  | Rural | 324/406 (79.8%) |
|  | Urban | 82/406 (20.2%) |
| **Health Facility** |  |  |
|  | Gishubi | 69/406 (17.0%) |
|  | Mahama | 66/406 (16.3%) |
|  | Masaka | 70/406 (17.2%) |
|  | Mushubati | 65/406 (16.0%) |
|  | Ndama | 68/406 (16.7%) |
|  | Tanda | 68/406 (16.7%) |
| **Sex** |  |  |
|  | Male | 178/406 (43.8%) |
|  | Female | 228/406 (56.2%) |
| **Age Categories** |  |  |
|  | <15 years | 65/406 (16%) |
|  | 15 years and above | 341/406 (84.0%) |
| **Occupation** |  |  |
|  | Farmer | 242/406 (59.6%) |
|  | Local Leader | 13/406 (3.2%) |
|  | Small Trade | 50/406 (12.3%) |
|  | Vocation work | 101/406 (25.0%) |
| **Religion** |  |  |
|  | Catholic | 163/406 (40.1%) |
|  | Protestant | 133/406 (32.8%) |
|  | Adventist | 55/406 (13.5%) |
|  | Evangelical | 55/406 (13.5%) |
| **Highest level of education** |  |  |
|  | No Formal Education | 85/406 (20.9%) |
|  | Primary | 204/406 (50.2%) |
|  | Secondary and above | 88/406 (28.8%) |

### Knowledge of Malaria Symptoms, Transmission, Control, Prevention, and Treatment

The study revealed that participants demonstrated a combined score of 81% (329/406) for understanding malaria signs and symptoms, with 99% (402/406) recognizing fever as a key symptom and 70.4% (286/406) identifying diarrhea as a possible indicator. Similarly, participants achieved a combined score of 72% (292/406) for awareness of malaria transmission, with all 100% (406/406) participants correctly identifying mosquito bites as the primary mode of transmission. However, significant misconceptions were noted, with rain exposure, immature sugarcane consumption, and bed bugs as causes of malaria by 40% (162/406), 35% (142/406), and 52% (211/406), respectively. Concerning treatment, 72% (292/406) of respondents were aware that artemisinin-based combination therapies (ACTs) are the recommended treatment for malaria, though 18.7% (76/406) expressed uncertainty about this. Preventive practices were encouraging, with 98% (389/406) agreeing that sleeping under treated nets is the most effective prevention strategy, 71.4% (290/406) endorsing indoor residual spraying, and 67% (272/406) supporting presumptive treatment. Overall, participants demonstrated a strong knowledge and practices in malaria prevention and control, achieving an average score of 74.6% (303/406), though targeted education is needed to address persistent misconceptions and improve treatment awareness (**Figure 2**).

**Figure 2.**
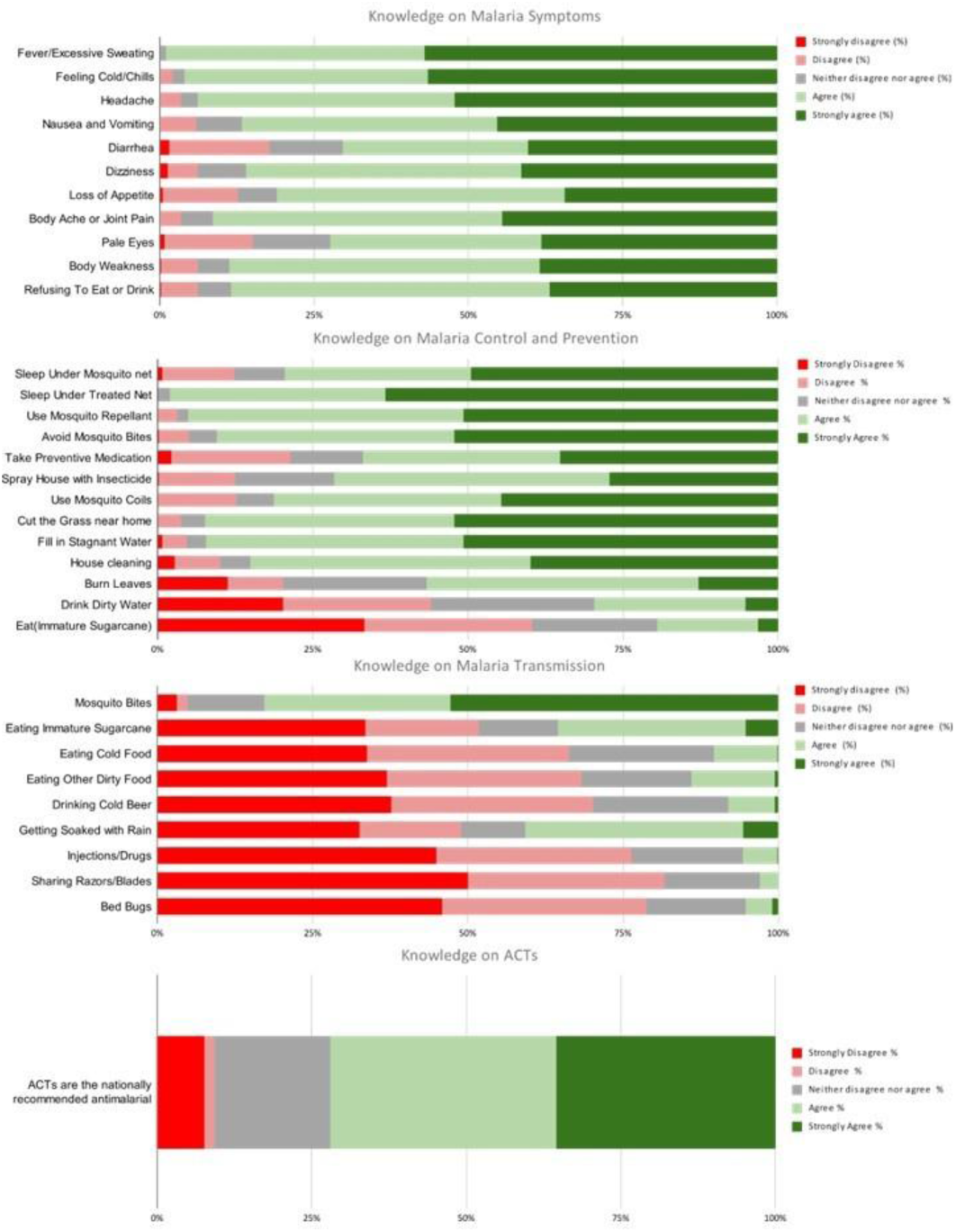
Details of knowledge on malaria symptoms, transmission, control and prevention and treatment. Red: strongly disagree; Pink: disagree; grey: neutral; Light green: agree; and Dark Green: Strongly Agree.

### Attitude on malaria control methods and treatment

Participants demonstrated mixed attitudes toward malaria control methods (Supplemental File 2 (**S2**). While 36.2% (147/406) expressed a positive view of chemicals used for indoor residual spraying (IRS), 26.6% (113/406) believed that spraying alone negates the need for other preventive measures. Additionally, 52.9% (215/406) agreed that sleeping under mosquito nets is sufficient to prevent malaria, and 76.3% (310/406) had a positive perception of the nets’ safety. Concerning net usability, 40.6% (165/406) approved of their use because they are provided for free, though a small proportion 5.6% (23/406) reported using nets for agricultural purposes. Regarding malaria treatment, most participants, 96.8% (393/406) sought care promptly when experiencing fever, and 68.7% (279/406) had a favorable attitude toward artemisinin-based combination therapies (ACTs). However, 20% (81/406) expressed a dislike for ACTs, and 8.6% (35/406) admitted to stopping treatment prematurely when they felt better (**Figure 3**).

**Figure 3.**
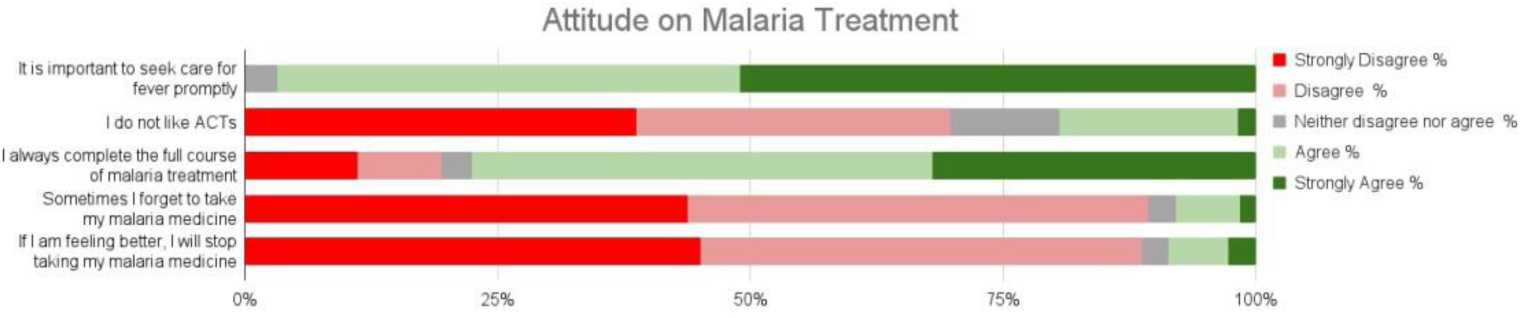
Malaria Treatment Attitudes and Reported Practices. Respondents’ degree of agreement with statements with regard to malaria treatment. Red: strongly disagree; Pink: disagree; grey: neutral; Light green: agree; and Dark Green: Strongly Agree.

### Malaria Treatment Practice

#### Adherence to malaria treatment

Of the 406 participants interviewed, 44.3% (180/406) had been diagnosed and treated for malaria in the last six months (diagnosed malaria). The ACT artemether-lumefantrine was used as malaria treatment for all patients (**Table 2**). Most study participants had been treated at the community level by CHWs 71.6% (129/180). Fewer received care at health care structures such as health centers 15.0% (27/180), outlets/pharmacies 11.1% (20/180) or via self-medication 2.2% (4/180). Only 46.1% (83/180) of the study participants diagnosed with malaria took the full three-day course of antimalarials. Additionally, 40.4% (73/180) of respondents took medication for less than the recommended period (<3 days), 10.5% (19/180) took medication for 4+ days, and for 2.7% (5/180) treatment duration was unknown.

**Table 2.** Adherence to malaria treatment (n=406)

|  | Diagnosed Malaria<br>n= 180(%) | Fever<br>n= 109(%) |
| --- | --- | --- |
| **Took any antimalarial in the last six months** |  |  |
| Yes | 180/406 (44.3%) | 109/406 (26.8%) |
| No | 226/406 (55.7%) | 292/406 (71.9%) |
| Unknown | 0/406 (0.0%) | 5/406 (1.2%) |
| **Antimalarial type** |  |  |
| artemether-lumefantrine (AL) | 180/180(100%) | 109/109 (100%) |
| **Source of antimalarial** |  |  |
| Health Center | 27/180 (15.0%) | 2/109 (1.8%) |
| Outlet/Pharmacy | 20/180 (11.1%) | 59/109 (54.1%) |
| Community Health Workers | 129/180 (71.6%) | 42/109 (38.5%) |
| Self-treatment | 4/180 (2.2%) | 6/109 (5.5%) |
| **Duration of treatment** |  |  |
| 1 Day | 7/180 (3.8%) | 1/109 (0.9%) |
| 2 Days | 66/180 (36.6%) | 26/109 (23.8%) |
| 3 Days | 83/180 (46.1%) | 73/109 (66.9%) |
| 4+ Days | 19/180 (10.5%) | 9/109 (8.2%) |
| Unknown | 5/180 (2.7%) | 0/109 (0.0%) |

A quarter of participants 26.8% (109/406) also reported taking an antimalarial for fever (without confirmatory diagnosis). The most common source of antimalarial drugs for fever were outlets/pharmacies, accounting for 54.1% (59/109) (**Table 2**). Two-thirds of participants 66.9% (73/109) took the medication as prescribed for three days, whereas 0.9% (1/109), 23.8% (26/109), and 8.2% (9/109) took the medication in one, two, and 4+ days, respectively.

#### Factors influencing adherence to malaria treatment

Factors associated with adherence to malaria treatment are presented in Table 3. Overall, adherence was associated with gender, education, occupation, location (urban), religion, and study site (health facility). In an adjusted multivariate analysis, only occupation and religion were found to be significantly associated with adherence to treatment among individuals diagnosed with malaria (**Table 3**). Vocational workers, including drivers, were less likely to be adherent compared to farmers (aOR=0.19; 95%CI:0.05-0.72; pvalue=0.015). Likewise, the adjusted odds of treatment adherence were significantly lower in participants who are Adventist compared to Catholics (aOR=0.10; 95%CI:0.02-0.49; pvalue=0.004). Females had twice the adjusted odds of adherence to AL compared to males (aOR=2.01; 95%CI:0.95-4.24; pvalue=0.067), but the result was not statistically significant. Having a history of usually completing treatment, stopping medications when feeling better or forgetting to take malaria medication was not associated with treatment adherence. However, participants who reported disliking ACT were less likely to be adherent to treatment; however, the finding was no longer significant after adjusting for other factors.

**Table 3.**
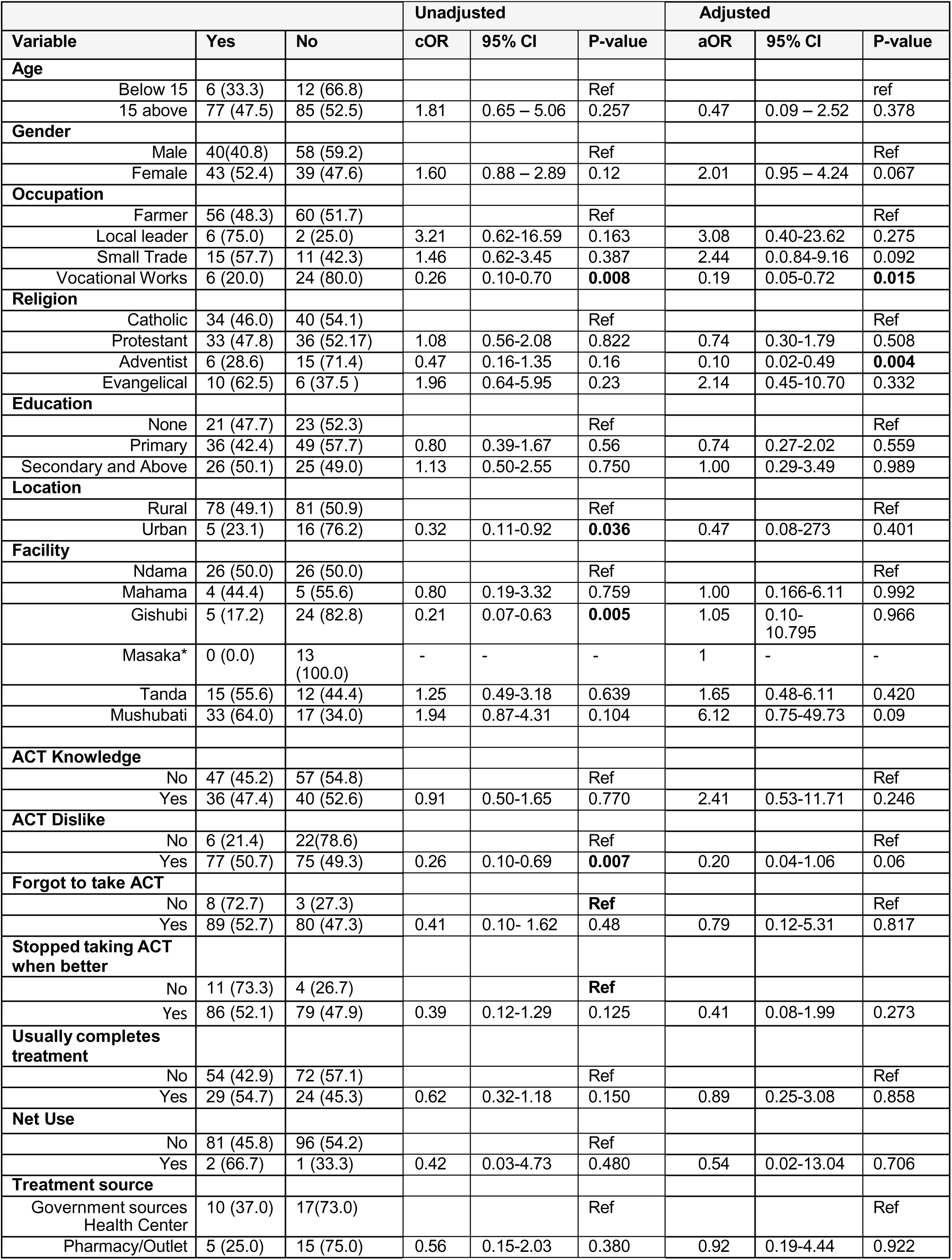
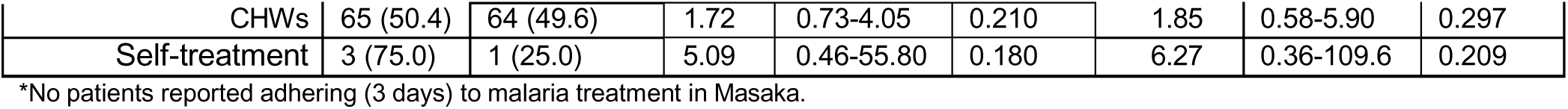
Factors associated with adherence to treatment for confirmed malaria diagnosis.

#### Factors influencing adherence to fever treatment

In an adjusted multivariate analysis, only religion was found to be significantly associated with adherence to fever treatment (**Table 4**). Protestants had four times the adjusted odds of adhering to fever treatment compared to Catholics (aOR=4.6; 95%CI: 1.02-20.8; p-value=0.05. Similarly, being Adventists was strongly associated with higher treatment adherence for fever (aOR=13.42; 95%CI: 1.45-125; p-value=0.02).

**Table 4.** Factors associated with antimalarial treatment adherence taken for fever (unconfirmed malaria)

| Variable |  |  | Unadjusted |  |  | Adjusted |  |  |
| --- | --- | --- | --- | --- | --- | --- | --- | --- |
|  | Yes | No | cOR | 95% CI | P-value | aOR | 95% CI | P-value |
| **Age** |  |  |  |  |  |  |  |  |
| Below 15 | 3 (27.3) | 8 (72.7) |  |  | Ref |  |  | Ref |
| 15 above | 34 (34.7) | 64 (65.3) | 0.70 | 0.18 - 2.8 | 0.62 | 0.60 | 0.08 - 4.38 | 0.61 |
| **Gender** |  |  |  |  |  |  |  |  |
| Male | 18 (33.3) | 36 (66.7) |  |  | Ref |  |  | Ref |
| Female | 19 (34.6) | 36 (65.5) | 0.95 | 0.43 - 2.1 | 0.89 | 0.68 | 0.23 - 2.00 | 0.49 |
| **Occupation** |  |  |  |  |  |  |  |  |
| Farmer | 18 (30.0) | 42 (70.0) |  |  | Ref |  |  | Ref |
| Local leader | 1 (33.3) | 2 (66.7) | 0.86 | 0.07 - 10.1 | 0.90 | 0.58 | 0.01 - 27.60 | 0.78 |
| Small Trade | 11 (52.4) | 10 (47.6) | 0.39 | 0.14 - 1.1 | 0.07 | 0.18 | 0.03 - 1.20 | 0.08 |
| Vocational Works | 7 (28.0) | 18 (72.0) | 1.10 | 0.39 - 3.1 | 0.85 | 1.71 | 0.33 - 8.82 | 0.52 |
| **Religion** |  |  |  |  |  |  |  |  |
| Catholic | 17 (37.8) | 28 (62.2) |  |  | Ref |  |  | Ref |
| Protestant | 10 (33.3) | 20 (66.7) | 1.21 | 0.46 - 3.2 | 0.70 | 5.68 | 0.78 - 41.39 | 0.086 |
| Adventist | 4 (28.6) | 10 (71.4) | 1.52 | 0.41 - 5.6 | 0.53 | 6.50 | 0.02 - 23.93 | 0.534 |
| Evangelical | 6(30.0) | 14 (70) | 1.41 | 0.45 - 4.4 | 0.55 | 2.25 | 0.44 - 11.37 | 0.323 |
| **Education** |  |  |  |  |  |  |  |  |
| None | 0 (0) | 10 (100) |  |  | Ref |  |  | Ref |
| Primary | 22 (41.5) | 31 (58.5) | 0.35 | 0.04 - 3.8 | 0.36 | 1.40 | 0.09-21.18 | 0.804 |
| Secondary and Above | 15 (32.6) | 31 (67.4) | 0.48 | 0.05 - 4.7 | Omitted | 1.522 | 0.10-22.79 | 0.762 |
| **Location** |  |  |  |  |  |  |  |  |
| Rural | 30 (36.6) | 52 (63.4) |  |  | Ref |  |  | Ref |
| Urban | 7 (25.9) | 20 (74.1) | 1.64 | 0.62 - 4.4 | 0.31 | 4.16 | 0.39 - 44.64 | 0.239 |
| **Facility\*** |  |  |  |  |  |  |  |  |
| Ndama | 1 (33.3) | 2 (66.7) |  |  | Ref |  |  | Ref |
| Mahama | 11 (73.3) | 4 (26.7) | 0.18 | 0.01 - 2.6 | 0.21 | 0.068 | 0.00 - 4.433 | 0.208 |
| Masaka | 6 (31.6) | 13 (68.4) | 1.08 | 0.08 -14.4 | 0.95 | 0.151 | 0.00 - 15.12 | 0.422 |
| Tanda | 8 (29.6) | 19 (70.4) | 1.19 | 0.09 - 15.0 | 0.89 | 0.504 | 0.00 - 24.45 | 0.730 |
| Mushubati | 11 (24.4) | 34 (75.6) | 1.55 | 0.13 - 18.7 | 0.73 | 1 | - | - |
| **ACT Knowledge** |  |  |  |  |  |  |  |  |
| No | 37 (61.7) | 23 (38.3) |  |  | Ref |  |  | Ref |
| Yes | 35 (71.4) | 14 (28.6) | 0.64 | 0.28 - 0.28 | 0.28 | 1.80 | 0.10- 30.35 | 0.683 |
| **ACT Dislike** |  |  |  |  |  |  |  |  |
| No | 3 (75.0) | 1 (25.0) |  |  | Ref |  |  | Ref |
| Yes | 69(65.7) | 36 (34.3) | 1.56 | 0.15 - 15.6 | 0.70 | 1.0 | - | - |
| **Forgot to take ACT\*\*** |  |  |  |  |  |  |  |  |
| No | 36 (34.6) | 68 (65.4) |  |  | Ref |  |  |  |
| Yes | 1 (20) | 4 (80) | 2.11 | 0.22-19.5 | 0.509 |  |  |  |
| **Stopped taking ACT when better\*\*** |  |  |  |  |  |  |  |  |
| No | 35 (34) | 68 (66) |  |  | Ref |  |  |  |
| Yes | 2 (33.3) | 4 (66.7) | 1.20 | 0.17-589 | 0.974 |  |  |  |
| **Usually complete Treatment** |  |  |  |  |  |  |  |  |
| No | 40 (59.7) | 27 (40.3) |  |  | Ref |  |  | Ref |
| Yes | 31(75.6) | 10 (24.4) | 0.47 | 0.20 - 1.1 | 0.09 | 0.47 | 0.03 - 6.81 | 0.582 |
| **Net Use** |  |  |  |  |  |  |  |  |
| No | 71(67.0) | 35 (33.0) |  |  | Ref |  |  | Ref |
| Yes | 1 (33.3) | 2 (66.7) | 4.05 | 0.35 - 46.2 | 0.26 | 1.0 | - | - |
| **Treatment Source\*\*** |  |  |  |  |  |  |  |  |
| Health Center | 0 (0.0) | 2 (100) |  |  | Ref |  |  | Ref |
| Pharmacy/Outlet | 25 (59.5) | 17 (40.9) | 0.29 | 0.03 - 2.7 | 0.28 | 0.50 | 0.04 - 7.15 | 0.61 |
| CHWs | 42 (71.2) | 17(28.8) | 0.49 | 0.05 - 4.5 | 0.53 | 0.14 | 0.01 - 2.83 | 0.17 |
| Self-treatment | 5 (83.3) | 1 (16.7) |  |  | - |  |  | - |

\*Gishubi was not included in this analysis as no patients from Gishubi reported seeking care for fever
\*\* The covariates "Forgot to take ACT," "Stopped taking ACT when better," and the "self-treatment" category under "Treatment Source" were not included in the adjusted analysis as fewer than 10 participants reported these practices a.

## Discussion

This study assessed knowledge, attitudes, and practices regarding malaria treatment among febrile patients and/or their caregivers visiting six government health facilities in Rwanda. The study revealed that participants demonstrated a high level of knowledge regarding malaria symptoms (99%), transmission (81%), prevention and control (74.6%), and treatment (72%). However, despite relatively high malaria knowledge, only 46.1% (83/180) of individuals diagnosed and treated for malaria completed the full dose of treatments within the recommended three days.

Patient adherence to antimalarials in Africa has been previously reported to be below 50%,^22^ and adherence to ACT ranges from 39% to 100%.^23^ To date, there is no published study on antimalarial adherence in Rwanda; however, the majority of adherence studies have taken place in neighboring countries. In Kenya, adherence to ACTs was reported to be only 29.4%.^24^ In contrast, another study conducted in Kenya revealed similar levels of patient adherence to ACT (64.1%).^25^ In Uganda, a study revealed good adherence at 69.7%.^26^ Similarly, ACT adherence was also reported to be 69.8% in Tanzania.^27^

In the review conducted by Bruxvoort et al, adherence was reported to be higher in studies where a diagnostic test was administered.^22^ In this study, 26.8% (109/406) of participants received anti-malarial drugs due to any fever without a confirmatory test. This is similar to observations in countries where malaria treatment is based on fever alone is commonly practiced. For instance, in Uganda, 34.3% of participants under five years old took anti-malarial drugs before getting tested.^28^ The study also revealed that 54.1% (59/109) of participants acquired anti-malarial drugs from outlets or pharmacies not affiliated with public health facilities. Similar trends were observed in a study on community pharmacies in Rwanda, which found that 88.5% of pharmacists were approached by patients seeking antimalarials, and 54% dispensed them without a prescription.^29^ This pattern is also evident in other African countries like Nigeria, where 46.5% of anti-malarial drugs are self-distributed by patients, and 35.8% are dispensed by outlets.^30^ Impressively, study findings revealed that all malaria cases, whether treated based on fever symptoms or a malaria diagnosis, were managed with AL (100%), regardless of the source of antimalarials. This aligns with Rwanda’s national malaria treatment guidelines, which designate AL as the primary treatment option nationwide for uncomplicated malaria^31^ Furthermore, the strict regulatory oversight by the Rwanda Food and Drugs Authority (Rwanda FDA) ensures the traceability and quality control of antimalarial medicines available on the market.^32^

Malaria treatment practices are crucial to successful malaria control programs. This study highlights a paradox in the Knowledge, Attitudes, and Practices (KAP) regarding malaria treatment in Rwanda. Despite high levels of knowledge and positive attitudes towards malaria treatment and prevention strategies, these do not seem to translate into corresponding behaviors, particularly in terms of adherence to antimalarial treatment. This discrepancy is concerning, as poor adherence to malaria treatment can potentiate the emergence of drug resistance, a significant threat in the era of developing resistance.^33^ This is particularly true for ACTs, where treatment in the face of ArtR parasites requires full dosing to achieve appropriate levels of partner drugs that can kill surviving parasites. Given the current status of artemisinin resistance in Rwanda and these findings that revealed poor adherence to ACT, there is an urgent need to study and better understand the drivers of poor adherence to ACT to identify solutions to improve medication-taking behaviors.

We found that individuals receiving treatment for fever were more likely to adhere to treatment compared to those with a malaria diagnosis (67% vs 46%). The difference observed in the two groups may be linked to the source of treatment. Participants with fever sought care from pharmacies/outlets, where as those with diagnosed malaria primarily sought treatment from CHW. Adherence to antimalarials have been reported to be higher for patients accessing treatment at public health facilities.^23,27^ CHWs are an extension of the health system and treatment adherence to medications source from them have been reported to be high (>90%), even when adding antibiotics to the treatment plan.^34^ Conversely, adherence to antimalarial treatment sourced at retail sector (outlets and pharmacies) was significantly lower compared to public health facilities (45% vs. 76%).^35^ Adherence behaviors may be influenced by patient-provider interactions during the consultation, such as detailed instructions from the health worker or the use of visual aids.^27,36^ However, patient adherence behaviors are not only influenced by patient and health worker characteristics, but can also be influenced by larger health system constraints such as stock outs, long wait-times, and patient loads.^37,38^ Rwanda aims to expand the health workforce by 2028, offering hope, that systemic barriers to care are improved, however this study highlights the importance of ensuring that both public and private facilities are equipped to provide patients with information on the importance of treatment adherence to patients.^39,40^

### Strengths and Limitations

To the best of our knowledge, this is the first study conducted in Rwanda to assess malaria treatment knowledge, attitudes, and adherence practices. The present findings are valuable data for NMCP and will be used as baseline information for monitoring Malaria treatment practices at the community level. However, this study has a number of limitations. First, the generalizability of this study is limited due to the small number of participants involved at only six health facilities, which may not be representative of other health facilities in Rwanda or other countries. Additionally, the small sample size limited our precision and ability to detect factors significantly associated with treatment adherence. However, despite the limited scale of this study, the data generated still provides important insights into community knowledge and malaria treatment practices and informs future investigations measuring malaria treatment adherence. Second, adherence was measured using self-report, which can be subject to recall and social desirability bias. To minimize bias, we used standardized survey tools based on Malaria Indicator Surveys and conducted interviews in a non-judgmental manner. Finally, participants were asked about their most recent fever episodes in the last six months, but the time when the event occurred was not recorded. This limitation prevents us from determining whether multiple reported cases refer to distinct episodes or potential double-counting of the same illness. This lack of temporal data may introduce recall bias and affect the accuracy of our incidence estimates. However, even without precise timing data, the results provide us with important information on the estimated level of antimalarial treatment adherence. Additionally, demographic, behavioral, and knowledge factors associated with adherence can be explored to identify areas or populations that may be targeted for interventions.

## Conclusions

This study revealed adequate knowledge of malaria, treatment options, and existing prevention strategies in selected health facilities in Rwanda. However, knowledge of malaria treatment did not translate to treatment-taking behaviors. Poor adherence to malaria treatment (<50%) was noted across all health facilities. With the emergence of antimalarial drug resistance in Rwanda, there is an urgent need to strengthen malaria case management practices and monitor adherence to antimalarial treatment.

## Data Availability

All data produced in the present study are available upon reasonable request to the authors

## Acknowledgments

The authors extend their thanks to all participants involved in the study and to the health facility administration for facilitating its implementation.

## Authors’ contributions

PG & KB contributed to the design of the work, data collection, analysis, interpretation of results, drafting, and revision of the manuscript. ES and PGI contributed to the study design and data analysis. NYW, JBM, and CK contributed to the design of the work and revision of the manuscript. JJJ, AJ, and JAB contributed to the design of the work, interpretation of results, and revision of the manuscript.

## Supplementary files

**Supplementary file 1 (S1): [Rwanda_KAP Gashema](https://drive.google.com/drive/u/0/folders/1ajh2ynjUH2KY9ks3m5abir0kVtHPmKCj)**

**Supplementary file 2 (S2): Malaria Prevention Attitudes**

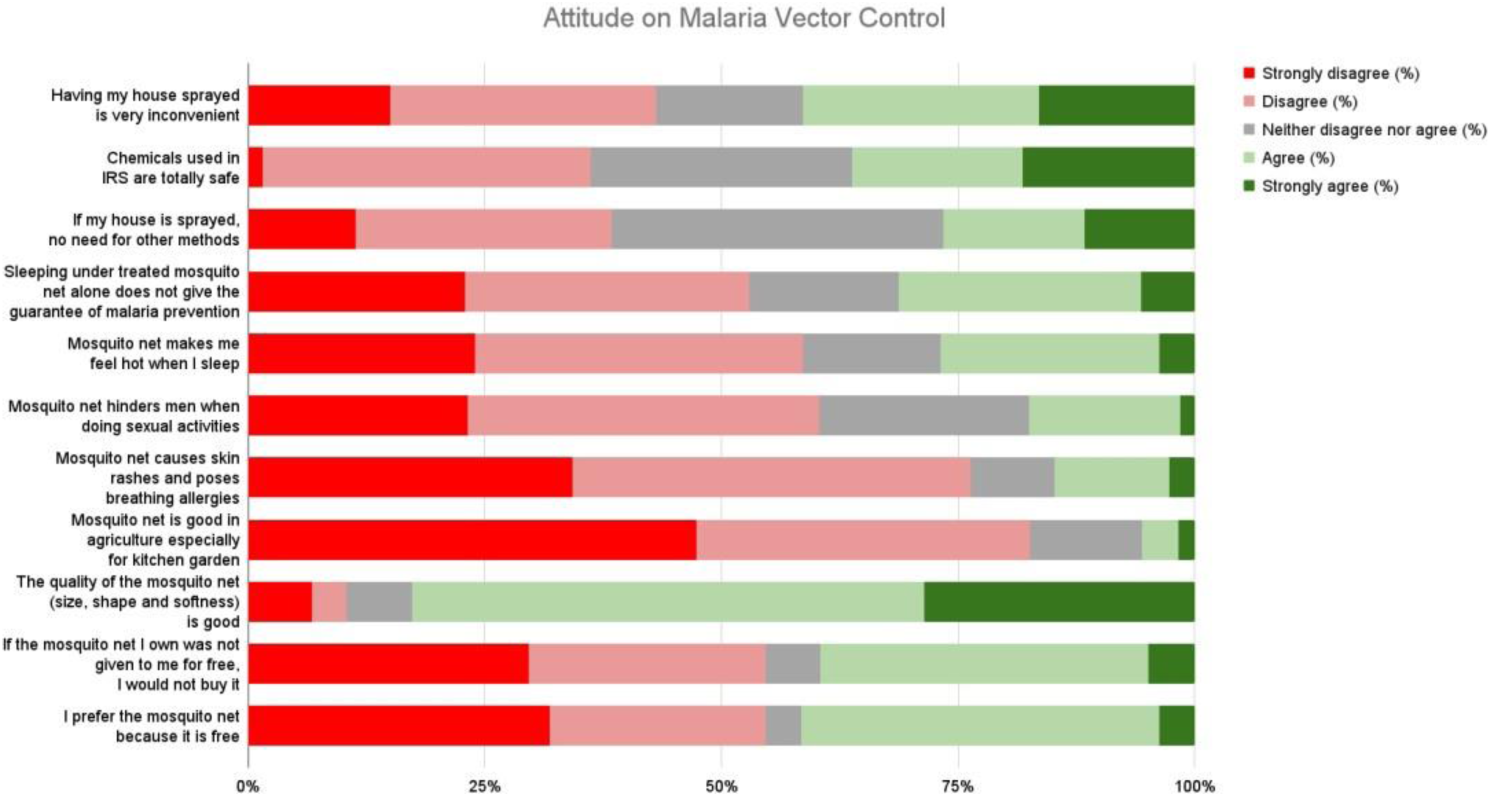

